# Plasma endostatin is an early creatinine independent predictor of acute kidney injury and need for renal replacement therapy in critical care

**DOI:** 10.1101/2024.04.25.24306345

**Authors:** Hazem Koozi, Jonas Engström, Martin Spångfors, Hans Friberg, Attila Frigyesi

## Abstract

**Purpose:** Endostatin is a promising biomarker for predicting acute kidney injury (AKI) and mortality in the intensive care unit (ICU). We investigated plasma endostatin upon ICU admission as a predictor of AKI, renal replacement therapy (RRT), and 30-day mortality.

**Methods:** A retrospective multicenter study was performed with admissions (ICU length of stay ≥24 hours) to four ICUs. KDIGO criteria defined AKI. Endostatin on ICU admission was compared to creatinine, cystatin C, and the Simplified Acute Physiology Score 3 (SAPS-3). Admissions with sepsis and creatinine <100 μmol/L on ICU admission underwent subgroup analyses. Regression models and the area under the receiver operating characteristic curve (AUC) were assessed.

**Results:** In total, 4449 admissions were included (43% sepsis and 61% AKI). Endostatin was associated with AKI (odds ratio [OR] 1.6, 95% confidence interval [CI] 1.4-1.7), future AKI (OR 1.5, 95% CI 1.4-1.7), future AKI stage 3 (OR 1.4, 95% CI 1.2-1.6), and RRT (OR 1.2, 95% CI 1.1-1.4) independently of creatinine and cystatin C, with similar results in sepsis. Endostatin was also associated with time to AKI (hazard ratio 1.2, 95% CI 1.1-1.2). For admissions with creatinine <100 μmol/L, endostatin (AUC 0.62, 95% CI 0.59-0.65) outperformed creatinine (AUC 0.51, 95% CI 0.49-0.54) and cystatin C (AUC 0.53, 95% CI 0.50-0.56) in predicting future AKI (p<0.001). Endostatin was not associated with 30-day mortality after adjusting for SAPS-3.

**Conclusion:** Endostatin is an early and potentially clinically useful biomarker for predicting AKI and RRT needs at ICU admission, especially in patients with low to mildly elevated creatinine.

## Introduction

Endostatin is a broad-spectrum angiogenesis inhibitor ^1,2^. It is a naturally occurring fragment of type XVIII collagen, an integral part of the basement membranes of renal tubular epithelium, Bowman’s capsule, mesangium, and renal capillaries ^3^. Beyond endostatin’s roles in cancer biology, studies have demonstrated that it is involved in inflammatory and cardiovascular diseases ^4–7^. Endostatin is also associated with prognosis in pulmonary arterial hypertension, heart failure, and in a general elderly population ^8–10^.

In recent studies, endostatin has shown potential as a marker of disease severity and prognosis in intensive care ^11,12^.

In the setting of critical illness, acute kidney injury (AKI) is a frequent complication associated with substantial morbidity and mortality ^13–15^. Sepsis is highly prevalent in the intensive care unit (ICU), accounts for a large proportion of AKI, and is linked with a worse prognosis than either syndrome alone ^16^. The clinically most established biomarker for AKI is creatinine. However, creatinine is a suboptimal indicator of AKI, as up to half of the glomerular filtration rate (GFR) may be lost before creatinine rises ^17^. Timely detection of AKI is crucial, as it facilitates prompt monitoring and could influence clinical decisions regarding hemodynamics, medications, and perioperative management. Novel biomarkers correlating with renal injury and/or function could play a vital role in this early detection ^18^.

Elevated plasma endostatin has been associated with chronic kidney disease (CKD) ^19,20^. Studies have demonstrated a correlation between endostatin levels and risk of AKI, but conflicting results have also been shown ^11,12,21,22^. Endostatin has been shown to predict renal recovery and mortality in AKI ^11,23^. Furthermore, endostatin has been associated with acute respiratory distress syndrome (ARDS) pathology and disease severity in COVID-19 ^24,25^. In the general intensive care population, a prospective study from 2017 concluded that endostatin has limited value as a predictor of mortality, while a more recent study showed that endostatin combined with age was useful in mortality prediction. Thus, the results regarding endostatin’s predictive value for mortality in the ICU have been inconsistent^12,22^. Additionally, most studies evaluating endostatin in intensive care have been relatively small.

The precise role and applicability of endostatin as a predictive biomarker in intensive care require further investigation in larger patient cohorts.

### Objectives

This study investigated the association between plasma endostatin levels upon ICU admission and AKI, renal replacement therapy (RRT), and 30-day mortality in critically ill patients. Endostatin was compared with more established biomarkers of AKI, creatinine and cystatin C, and the Simplified Acute Physiology Score (SAPS-3) upon ICU admission. Additionally, we performed a subgroup analysis in patients with sepsis.

## Methods

### Study design and setting

A retrospective multicenter cohort study was performed as part of the Swecrit project, described in detail by Frigyesi et al. ^26^. The Strengthening the Reporting of Observational Studies in Epidemiology (STROBE) guidelines were followed ^27^.

Admissions to one of four general ICUs in southern Sweden (Skåne University Hospital in Lund and Malmö, Helsingborg Hospital, and Kristianstad Hospital) between 2015 and 2018 were consecutively included.

### Participants

All ICU admissions were screened for inclusion. Transfers between participating ICUs were consolidated into single and unique admissions. Exclusion criteria comprised admissions with missing biobank samples, incorrect sampling, patients or their next of kin opting out from the study, ICU length of stay less than 24 hours, missing endostatin analysis, and missing data from the Patient Administrative System for Intensive Care Units (PASIVA, see below).

### Variables

Endostatin, creatinine, and cystatin C were analysed on ICU admission. Creatinine was also analysed on the first 2 days of the ICU stay or until ICU discharge, whichever occurred sooner. For baseline creatinine, the value closest to ICU admission within 7 to 365 days before was recorded. If baseline creatinine was missing, it was estimated using the 2021 Chronic Kidney Disease Epidemiology Collaboration (CKD-EPI) creatinine equation ^28^. Urine output was recorded on the two mornings following ICU admission or until ICU discharge. If urine output could not be monitored for 24 hours, it was extrapolated from hourly urine output. AKI was defined as fulfilment of KDIGO criteria within 2 days of ICU admission or RRT during the ICU stay ^29^. For admissions to be classified as not having AKI, no missing values in any creatinine and urine output parameters were accepted. For admissions to be classified as having AKI, missing values in some creatinine and/or urine output parameters were acceptable as long as they met the KDIGO criteria. RRT was regarded as AKI unless the admission had a diagnosis of dialysable intoxication, in which case they needed to fulfil another KDIGO criteria. For the assessment of early AKI, the outcome of future AKI was created. Future AKI was defined as the absence of AKI upon ICU admission, followed by its subsequent development. Future AKI stage 3 was defined as the absence of AKI stage 3 upon ICU admission, followed by its subsequent development.

For variables included in SAPS-3 and the Sequential Organ Failure Assessment (SOFA) (excluding admission creatinine), the worst recorded values within 1 hour of ICU admission were recorded ^30,31^. The value closest in time, within 24 hours, of ICU admission was recorded for C-reactive protein (CRP), lactate, and procalcitonin.

Sepsis was defined as a SOFA score of 2 or more upon ICU admission and suspicion of infection within 24 hours of ICU admission. The baseline SOFA score was assumed to be 0. Suspected infection was defined as blood culture sampling and concurrent administration of antibiotics within 24 hours before to 72 hours after blood culture sampling, as suggested by the Sepsis-3 task force ^31^. Septic shock was defined as vasopressor use and lactate exceeding 2 mmol/L at ICU admission. Immunosuppressive treatment, metastatic and haematological cancer, cirrhosis, and chronic heart failure were all defined according to SAPS-3 ^30^.

### Data sources

Endostatin, cystatin C, and admission creatinine were retrospectively batch analysed from prospectively collected blood samples. Blood samples were collected upon ICU admission using ethylenediamine tetraacetic acid (EDTA) vacutainers and centrifuged to obtain EDTA plasma. The plasma samples were aliquoted and stored in the SWECRIT biobank at −80°C. Samples had to be collected within 6 hours of ICU admission. In cases where the sampling time was missing, samples were included if the freezing time fell within the 6-hour time frame.

Endostatin analyses were performed using commercial sandwich kits (DY1098, R&D Systems, Minneapolis, MN, USA). A monoclonal antibody specific for endostatin was coated onto microtiter plates. The plates were blocked with bovine serum albumin and then washed, and samples and standards were added to the wells, after which the peptide was bound to the immobilised antibodies. A biotinylated endostatin-specific antibody was added after washing. A streptavidin-HR conjugate was added to the wells after incubation and washing, and a substrate solution was added after another incubation and washing cycle. The absorbance was measured in a SpectraMax 250 (Molecular Devices, Sunnyvale, CA, USA). Endostatin values were determined by comparing the optical density of the samples with the standard curve. All assays were calibrated against highly purified recombinant human endostatin. Measurements were performed blinded, without knowledge of the clinical diagnoses. The total coefficient of variation for the endostatin assay was approximately 6%.

Admission creatinine and cystatin C were analysed on a Mindray BS380 chemistry analyser (Mindray Medical International, Shenzhen, China) using IDMS traceable enzymatic creatinine reagents from Abbott Laboratories (Abbott Park, IL, USA) and particle-enhanced turbidimetric cystatin C reagents from Gentian (Moss, Norway).

Clinical data, including SAPS-3 and SOFA parameters, survival data, age, and sex, were automatically extracted from PASIVA. PASIVA is a digital system where treating clinicians submit data to the Swedish Intensive Care Registry (SIR) during patients’ ICU stay. Baseline creatinine was extracted from electronic medical records.

Comorbidities not included in SAPS-3, such as height and body weight, were mainly collected for admissions with sepsis using manual electronic medical records review. CRP, lactate, and procalcitonin levels were also mainly collected for admissions with sepsis using automatic extraction from electronic medical records.

### Study size

The sample size was determined by the number of ICU admissions, the validity of blood samples, the number of opt-outs, the ICU length of stay, and the number of admissions with missing endostatin analyses or PASIVA data.

### Bias

Neither treating clinicians nor data collectors had any knowledge of endostatin levels. Trained data collectors performed the manual data recording. Guidelines for data collection were standardised and precisely outlined. The management of missing data was discussed and decided on collectively in the study group.

### Statistics

All statistical analyses were performed in R^32^. The median value was calculated for general characteristics, SAPS-3, physiological parameters, and biochemistry. The mean value was calculated for the Glasgow Coma Scale (GCS) and SOFA score. The Wilcoxon rank-sum test or the Kruskal-Wallis test was used to compare differences in a continuous variable’s median. Pearson’s chi-squared test or the chi-square test of independence was used to compare proportions. The unpaired t-test or analysis of variance was used to compare means. Significance was determined as a p-value of less than 0.05.

Missing body weight was imputed based on age and gender using the standardised function in R.

Local polynomial regression was employed to show trends in the association between endostatin, outcomes, and creatinine ^33^.

Correlations between variables were calculated using pairwise Spearman rank correlations. A correlation network was then visualised to represent relationships among variables of interest.

Logistic regression was utilised to create prediction models of AKI, RRT, and 30-day mortality as binary outcomes. Cox proportional hazard was employed to assess time to AKI. To facilitate calculation and comparisons of odds ratios (ORs) and hazard ratios (HRs) in regression analyses, predictor variables underwent log transformation and z-normalisation. For AKI and RRT, adjusted ORs and HRs were reported for a model where all three predictor variables (endostatin, creatinine, and cystatin C) were included simultaneously. For 30-day mortality, adjusted ORs for endostatin were reported for a model where endostatin and SAPS-3 were included simultaneously.

Discrimination was evaluated using the area under the receiver operating characteristic (ROC) curve (AUC). Differences in AUC were assessed using the method proposed by DeLong et al. ^34^. The accuracy of prediction was assessed using the Brier score.

## Results

### Participants

A total of 8406 ICU admissions were identified. After exclusions due to missing or invalid biobank sampling and opt-outs, 7130 admissions remained. Of these, 2621 admissions had an ICU stay of less than 24 hours, 23 had missing endostatin analyses, and 9 lacked PASIVA data. See supplemental figure 1. Consequently, the final study population comprised 4449 ICU admissions, with 1907 (43%) classified as having sepsis.

### Descriptive data

#### Study population

Admissions with AKI were older, to a larger extent males, had a higher body mass index (BMI), and a higher proportion of sepsis and septic shock. They also had more comorbidities and higher SAPS-3 and SOFA scores. Their baseline creatinine, admission creatinine, and cystatin C were higher than those without AKI. See Table 1. The proportion of missing body weight was 59%.

**Table 1.** Characteristics of the study population. Values are medians with interquartile ranges unless otherwise specified. P-values were calculated using the Wilcoxon rank-sum test and Pearson’s chi-square test as appropriate unless otherwise specified. ^*^Presented as means with standard deviations, and p-value calculated using unpaired t-test. *AKI* acute kidney injury, *SAPS-3* Simplified Acute Physiology Score 3, *Pa0*_2_ arterial partial pressure of oxygen, *Fi0*_2_ fraction of inspired oxygen (%), *SOFA* Sequential Organ Failure Assessment, *NIV* non-invasive ventilation, *RRT* renal replacement therapy, *ICU* intensive care unit.

|  | All | No AKI | AKI | p-value | Missing (%) |
| --- | --- | --- | --- | --- | --- |
| Number (%) | 4449 (100) | 1067 (24) | 2724 (61) |  | 15 |
| Age (years) | 68 (56-75) | 64 (49-72) | 70 (60-76) | <0.001 | 0 |
| Male sex (%) | 60 | 58 | 62 | 0.017 | 0 |
| Body mass index (kg/m<sup>2</sup>) | 27 (23-31) | 25 (22-30) | 27 (24-32) | <0.001 | 68 |
| Sepsis (%) | 43 | 36 | 49 | <0.001 | 0 |
| Septic shock (%) | 19 | 11 | 25 | <0.001 | 0 |
| **Comorbidities** |  |  |  |  |  |
| Immunosuppressive therapy (%) | 8.5 | 5.9 | 10 | <0.001 | 0 |
| Metastatic cancer (%) | 11 | 13 | 11 | 0.050 | 0 |
| Haematological cancer (%) | 2.7 | 1.8 | 3.3 | 0.018 | 0 |
| Cirrhosis (%) | 2.0 | 0.75 | 2.8 | <0.001 | 0 |
| Chronic heart failure (%) | 10 | 5.5 | 13 | <0.001 | 0 |
| Hypertension (%) | 34 | 27 | 38 | <0.001 | 42 |
| Diabetes mellitus (%) | 25 | 15 | 27 | <0.001 | 55 |
| Chronic kidney disease (%) | 11 | 2.2 | 14 | <0.001 | 55 |
| Chronic dialysis (%) | 2.6 | 0 | 3.7 | <0.001 | 55 |
| **Organ dysfunction and illness severity** |  |  |  |  |  |
| SAPS-3 score | 63 (51-74) | 57 (46-67) | 67 (57-77) | <0.001 | 0 |
| SOFA score* | 6.9 (4.0) | 5.4 (3.3) | 8.0 (4.0) | <0.001 | 2.5 |
| PaO<sub>2</sub>/FiO<sub>2</sub> (kPa) | 28 (17-44) | 30 (18-46) | 27 (16-42) | <0.001 | 14 |
| Cardiovascular SOFA* | 2.1 (1.7) | 1.7 (1.7) | 2.4 (1.7) | <0.001 | 1.5 |
| Mean arterial pressure (mmHg) | 65 (59-77) | 70 (60-80) | 65 (55-73) | <0.001 | 2.4 |
| Glasgow Coma Scale* | 12 (4.3) | 12 (4.3) | 11 (4.3) | 0.036 | 1.4 |
| Body temperature (°C) | 37 (36-38) | 37 (36-38) | 37 (36-38) | <0.001 | 0.67 |
| **Biochemistry** |  |  |  |  |  |
| Plasma Endostatin (ng/mL) | 62 (45-88) | 49 (38-63) | 73 (53-100) | <0.001 | 0 |
| Baseline creatinine (μmol/L) | 79 (62-110) | 70 (58-88) | 85 (58-88) | <0.001 | 34 |
| Creatinine (μmol/L) | 98 (68-150) | 72 (55-89) | 130 (93-210) | <0.001 | 0.022 |
| Cystatin C (mg/L) | 1.2 (0.75-1.9) | 0.79 (0.61-1.1) | 1.6 (1.0-2.4) | <0.001 | 0.022 |
| White blood cell count (×10<sup>9</sup>/L) | 13 (8.8-18) | 12.8 (8.8-17) | 13.5 (8.9-19) | 0.0080 | 5.1 |
| Platelet count (×10<sup>9</sup>/L) | 210 (150-290) | 230 (170-300) | 200 (140-280) | <0.001 | 5.7 |
| Lactate (mmol/L) | 2.0 (1.1-3.8) | 1.5 (1.0-2.7) | 2.4 (1.3-4.7) | <0.001 | 0.25 |
| C-reactive protein (mg/L) | 44 (7.4-150) | 42 (4.9-140) | 58 (12-170) | <0.001 | 0.65 |
| Procalcitonin (μg/L) | 1.8 (0.39-11) | 0.60 (0.21-2.8) | 3.3 (0.69-20) | <0.001 | 52 |
| Bilirubin (μmol/L) | 10 (6.0-17) | 9.0 (6.0-14) | 11 (7.0-19) | <0.001 | 6.4 |
| **Outcomes** |  |  |  |  |  |
| Mechanical ventilation/NIV (%) | 61 | 64 | 62 | 0.21 | 8.5 |
| Vasopressor therapy (%) | 55 | 48 | 64 | <0.001 | 0 |
| RRT (%) | 14 | 0.28 | 22 | <0.001 | 0 |
| ICU length of stay (days) | 2.7 (1.7-4.6) | 3.0 (2.0-5.2) | 3.2 (1.9-6.0) | 0.16 | 0 |
| 30-day mortality (%) | 26 | 19 | 32 | <0.001 | 0.022 |

Admissions with higher endostatin were older, had higher BMI, higher rates of sepsis and septic shock, and higher rates of almost all comorbidities. They also had higher SAPS-3 and SOFA scores and higher degrees of organ dysfunction, except for neurologic dysfunction. Admissions with higher endostatin had higher creatinine, cystatin C, lactate, CRP, and procalcitonin. See Table 2.

**Table 2.** Characteristics of the study population divided into plasma endostatin (ng/mL) tertiles. Values are medians with interquartile ranges unless otherwise specified. P-values were calculated using the Kruskal-Wallis test and the chi-square test of independence as appropriate unless otherwise specified. ^*^Presented as means with standard deviations, and p-value calculated using analysis of variance. *SAPS-3* Simplified Acute Physiology Score 3, *PaO*_*2*_ arterial partial pressure of oxygen, *FiO*_*2*_ fraction of inspired oxygen (%), *SOFA* Sequential Organ Failure Assessment, *NIV* non-invasive ventilation, *AKI* acute kidney injury, *RRT* renal replacement therapy, *ICU* intensive care unit.

|  | Endostatin<50 | Endostatin 50-77 | Endostatin>77 | p-value |
| --- | --- | --- | --- | --- |
| Number (%) | 1472 (33) | 1500 (34) | 1477 (33) |  |
| Age (years) | 61 (44-71) | 69 (60-76) | 71 (63-77) | <0.001 |
| Male sex (%) | 61 | 60 | 60 | 0.76 |
| Body mass index (kg/m<sup>2</sup>) | 25 (22-30) | 26 (23-31) | 27 (24-32) | <0.001 |
| Sepsis (%) | 33 | 43 | 53 | <0.001 |
| Septic shock (%) | 12 | 19 | 24 | <0.001 |
| **Comorbidities** |  |  |  |  |
| Immunosuppressive therapy (%) | 6.7 | 8.7 | 10.2 | 0.0024 |
| Metastatic cancer (%) | 11 | 12 | 11 | 0.45 |
| Haematological cancer (%) | 1.8 | 3.3 | 3.1 | 0.036 |
| Cirrhosis (%) | 2.8 | 1.3 | 2.0 | 0.020 |
| Chronic heart failure (%) | 4.1 | 8.5 | 19 | <0.001 |
| Hypertension (%) | 24 | 38 | 39 | <0.001 |
| Diabetes mellitus (%) | 14 | 24 | 32 | <0.001 |
| Chronic kidney disease (%) | 2.6 | 4.5 | 23 | <0.001 |
| Chronic dialysis (%) | 0.39 | 0.43 | 5.9 | <0.001 |
| **Organ dysfunction and illness severity** |  |  |  |  |
| SAPS-3 score | 56 (45-66) | 63 (52-73) | 69 (59-79) | <0.001 |
| SOFA score* | 5.6 (3.7) | 6.7 (3.9) | 8.2 (4.0) | <0.001 |
| PaO<sub>2</sub>/FiO<sub>2</sub> (kPa) | 33 (19-49) | 27 (16-41) | 26 (16-41) | <0.001 |
| Cardiovascular SOFA* | 1.8 (1.7) | 2.1 (1.7) | 2.3 (1.7) | <0.001 |
| Mean arterial pressure (mmHg) | 68 (60-80) | 65 (60-80) | 65 (54-75) | <0.001 |
| Glasgow Coma Scale* | 12 (4.4) | 12 (4.4) | 11 (4.2) | 0.96 |
| Body temperature (°C) | 37 (36-38) | 37 (36-38) | 37 (36-38) | 0.0013 |
| **Biochemistry** |  |  |  |  |
| Baseline creatinine (μmol/L) | 69 (56-81) | 76 (60-94) | 110 (76-160) | <0.001 |
| Creatinine (μmol/L) | 74 (57-96) | 93 (69-130) | 170 (110-290) | <0.001 |
| Cystatin C (mg/L) | 0.74 (0.58-1.0) | 1.1 (0.82-1.6) | 2.1 (1.5-3.0) | <0.001 |
| White blood cell count (×10<sup>9</sup>/L) | 12 (8.0-17) | 13 (9.2-19) | 14 (9.3-19) | <0.001 |
| Platelet count (×10<sup>9</sup>/L) | 200 (150-300) | 220 (150-300) | 220 (140-300) | <0.001 |
| Lactate (mmol/L) | 1.8 (1.1-3.3) | 2.0 (1.2-3.7) | 2.2 (1.2-4.5) | <0.001 |
| C-reactive protein (mg/L) | 17 (2.4-95) | 45 (7.2-140) | 77 (21-200) | <0.001 |
| Procalcitonin (μg/L) | 0.81 (0.23-5.7) | 1.4 (0.31-9.0) | 3.2 (0.78-21) | <0.001 |
| Bilirubin (μmol/L) | 10 (6.0-16) | 10 (6.0-17) | 10 (6.0-17) | 0.41 |
| **Outcomes** |  |  |  |  |
| Mechanical ventilation/NIV (%) | 56 | 58 | 55 | 0.20 |
| Vasopressor therapy (%) | 47 | 57 | 62 | <0.001 |
| AKI (%) | 38 | 58 | 83 | <0.001 |
| AKI ICU admission (%) | 18 | 35 | 63 | <0.001 |
| AKI ICU day 1 (%) | 27 | 46 | 77 | <0.001 |
| AKI ICU day 2 (%) | 21 | 35 | 62 | <0.001 |
| RRT (%) | 4.1 | 9.3 | 28 | <0.001 |
| ICU length of stay (days) | 2.2 (1.6-4.3) | 2.7 (1.7-5.1) | 3.2 (1.9-5.7) | <0.001 |
| 30-day mortality (%) | 18 | 26 | 34 | <0.001 |

Admissions with a creatinine <100 μmol/L upon ICU admission comprised 52% of the study population.

#### Sepsis subgroup

The rate of septic shock was 53%. Sepsis admissions with AKI were older and had higher BMI. Most comorbidities were more common in admissions with AKI. Admissions with AKI had higher SOFA and SAPS-3 scores and higher degrees of dysfunction in most organ systems. Baseline creatinine, admission creatinine, and cystatin C were higher in admissions with AKI. AKI admissions also exhibited higher lactate, procalcitonin, and bilirubin levels. See supplemental table 1. Admissions with sepsis had higher median endostatin (67 ng/mL vs. 57 ng/mL, p<0.001), creatinine (110 vs. 105, p<0.001), and cystatin C (1.4 vs. 0.99, p<0.001) but not baseline creatinine (80 vs. 79, p=0.32) compared to admissions without sepsis. The rate of missing body weight was 26%.

### Outcomes

#### Study population

Mortality within 30 days was 26%, and higher in admissions with AKI (32%) than admissions without AKI (19%). The rate of AKI was 61%, and the rate of RRT was 14%. Admissions with AKI also had a higher rate of vasopressor therapy. ICU length of stay was the same for admissions with and without AKI. See Table 1. In those with AKI, 34% had AKI stage 1, 23% had stage 2, and 43% had stage 3.

Admissions with a creatinine <100 μmol/L upon ICU admission had an AKI rate of 48% AKI and an RRT rate of 2.7%.

#### Sepsis subgroup

In the sepsis subgroup, 30-day mortality was 28%, with higher mortality in admissions with AKI. The AKI rate was 72%, and 19% received RRT. Vasopressor therapy was more common in AKI. ICU length of stay was the same with and without AKI. See supplemental table 1. Admissions with sepsis had a higher rate of AKI (70% vs. 54%, p<0.001), RRT (19% vs. 9.8%, p<0.001), and 30-day mortality (28% vs. 24%, p=0.0019) compared to admissions without sepsis. Of those with AKI, 32% had AKI stage 1, 20% had stage 2, and 48% had stage 3.

### Relationship between endostatin and outcome

The relationships between endostatin and AKI, RRT, and 30-day mortality are illustrated in Figure 1. The relationships between endostatin, admission creatinine, and AKI are displayed in Figure 2. A correlation network involving endostatin, other biomarkers, AKI, RRT, sepsis, and age was constructed to visualise associations; see Figure 3.

**Figure 1.**
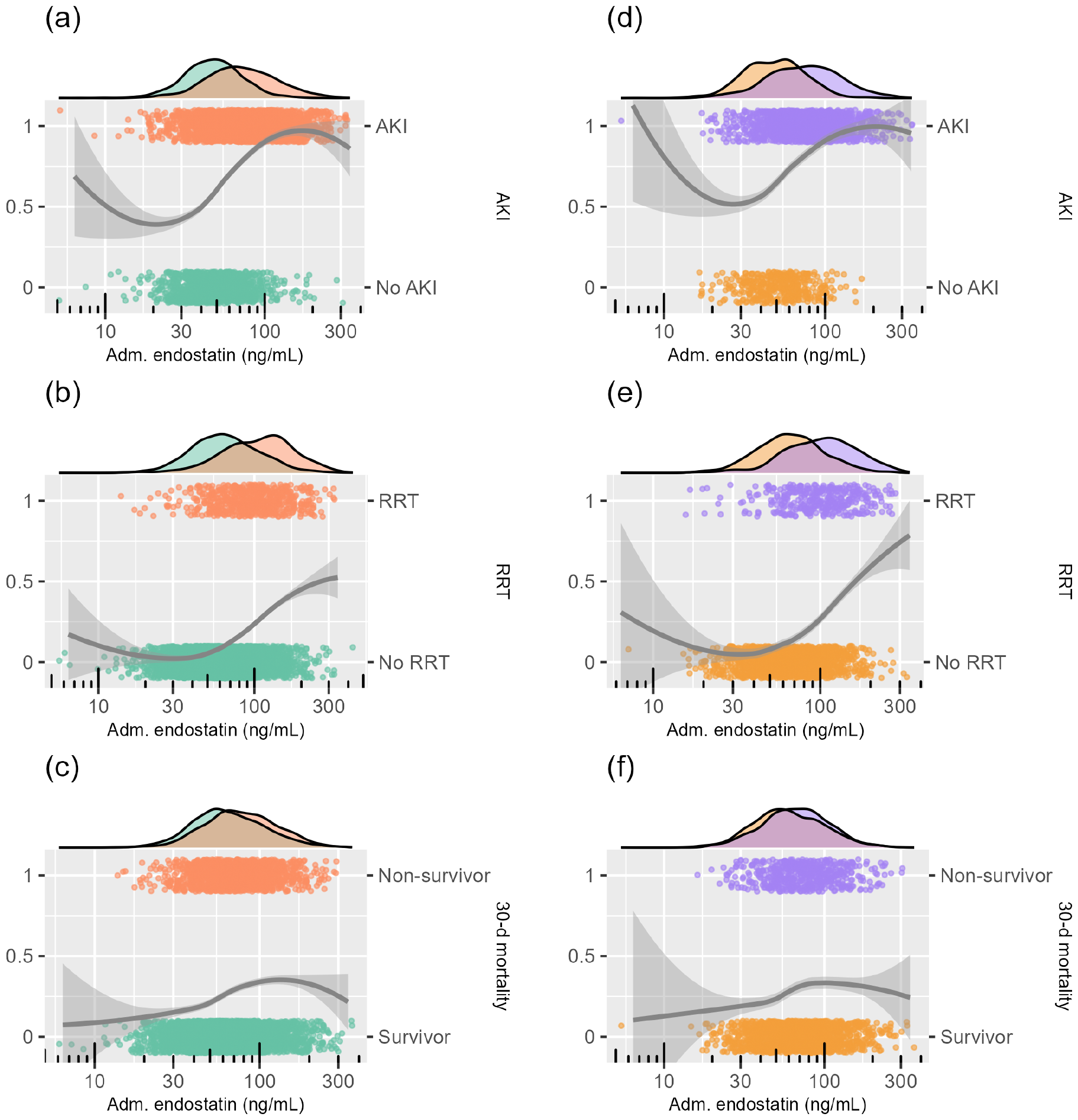
The relationship between admission plasma endostatin and AKI, RRT, and 30-day mortality in the study population (a-c) and the sepsis subgroup (d-f) visualised in scatter and density plots. The x-axes have logarithmic scales. The points show the outcome (AKI, RRT, or 30-day mortality) versus plasma endostatin level in individual patients. Continuous lines represent smoothed conditional means, while the accompanying shading indicates the corresponding 95% confidence intervals. *Adm* admission, *AKI* acute kidney injury, *RRT* renal replacement therapy, *30-d* 30-day.

**Figure 2.**
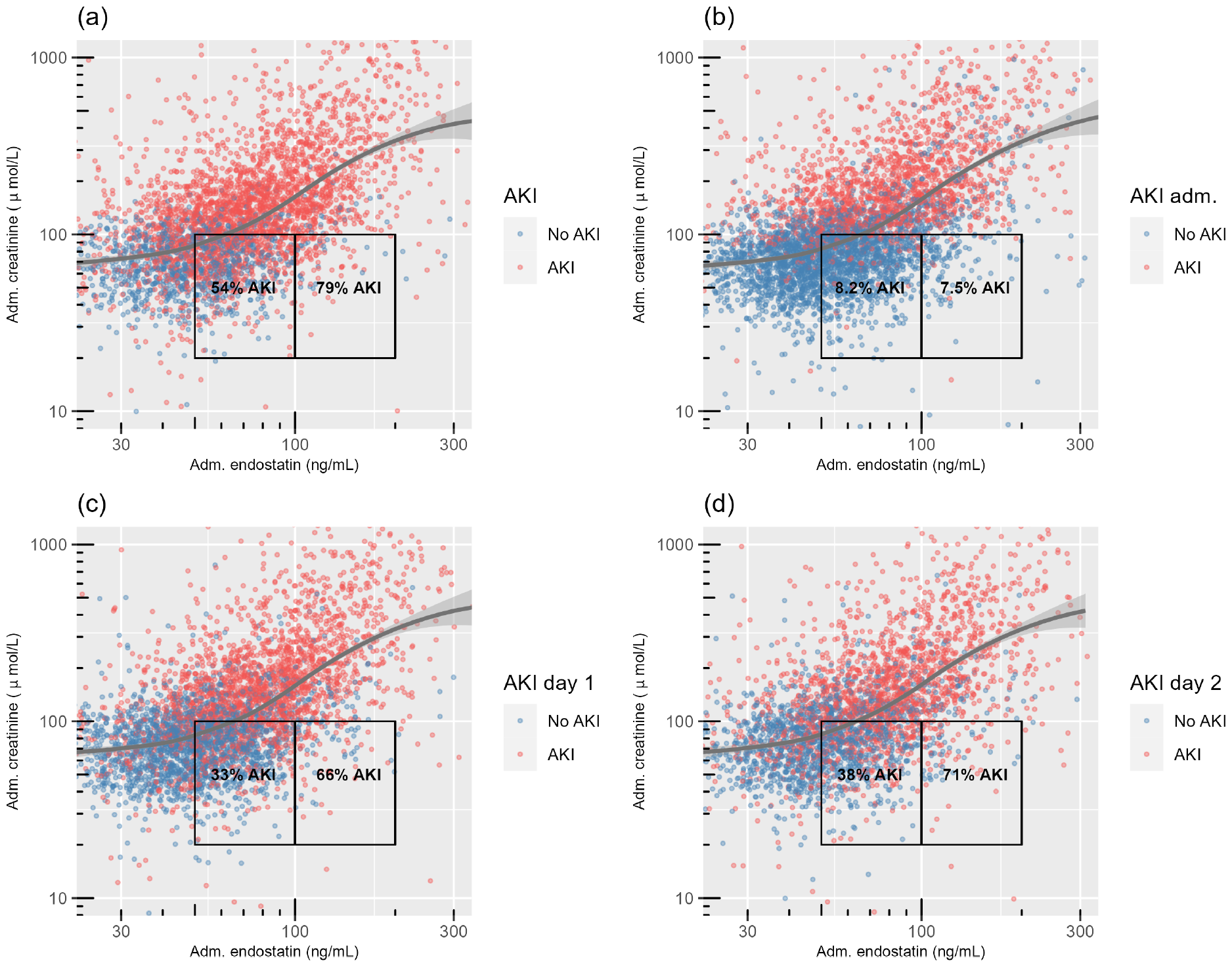
The relationship between admission plasma endostatin, admission plasma creatinine, and AKI in the study population visualised in scatter plots. The boxes to the left in each plot indicate ICU admissions with a creatinine of 20-100 μmol/L and endostatin of 50-100 ng/mL, while the right boxes indicate creatinine of 20-100 μmol/L and endostatin of 100-200 ng/mL. P-values for AKI rate comparison between boxes: <0.001 (a), 0.96 (b), <0.001 (c), <0.001 (d). P-values were calculated using Pearson’s chi-squared test. The y- and x-axes have logarithmic scales. Continuous lines show smoothed conditional means, while the accompanying shading indicates the corresponding 95% confidence intervals. *Adm* admission, *AKI* acute kidney injury, *ICU* intensive care unit.

**Figure 3.**
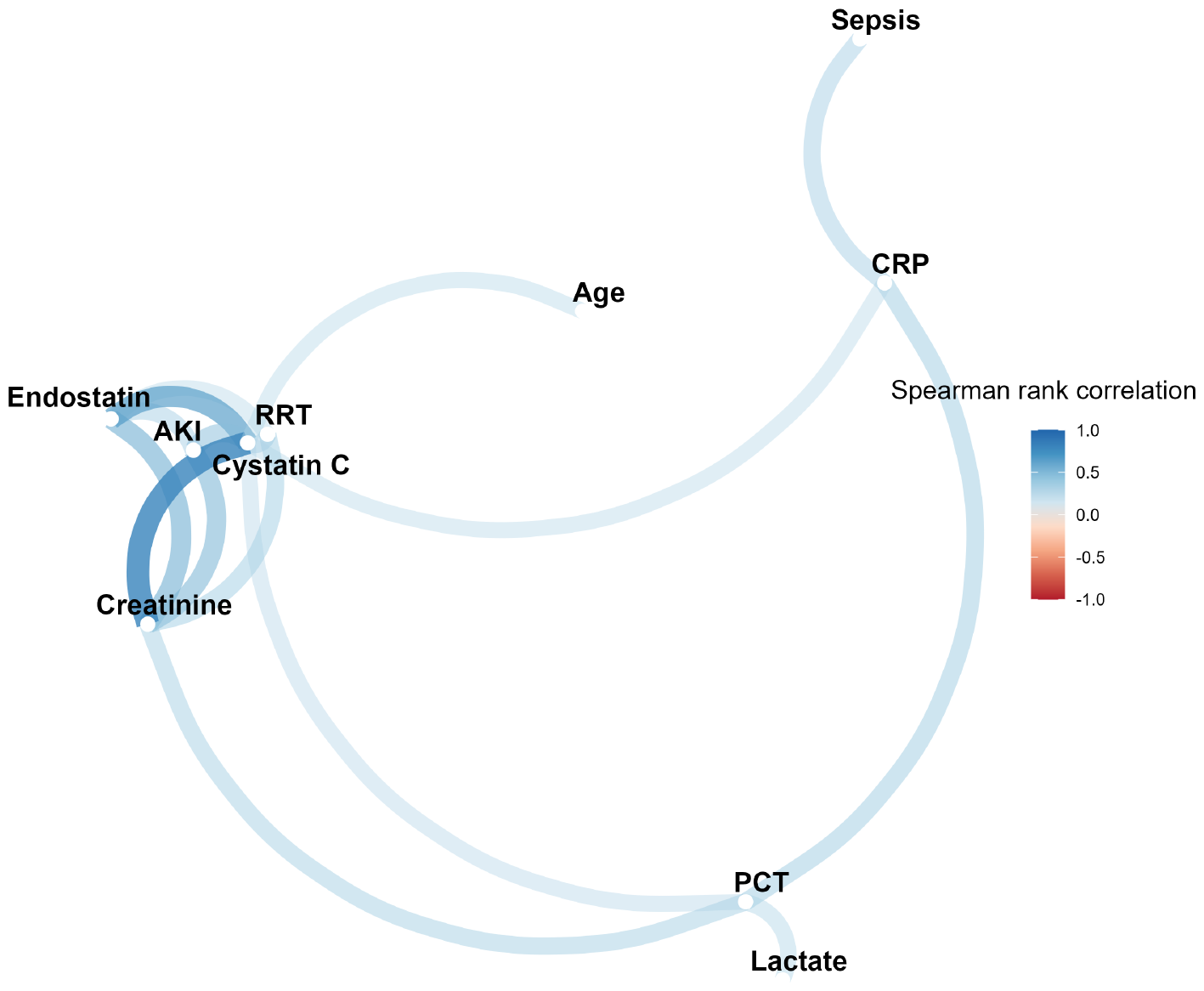
Correlation network for plasma endostatin, other biomarkers, AKI, RRT, sepsis, and age in the study population. The correlation network depicts pairwise relationships among variables, assigning a colour to edges based on the Spearman rank correlation coefficients. Colour intensity reflects the correlation strength, with darker shades indicating stronger associations. Only variables directly or indirectly linked to endostatin through absolute correlations exceeding 0.3 are displayed. The spatial arrangement of nodes groups variables into clusters with pronounced interconnections, highlighting strongly correlated variables. *AKI* acute kidney injury, *RRT* renal replacement therapy. *PCT* procalcitonin, *CRP* C-reactive protein.

### Main results

The AUCs and Brier scores of endostatin, creatinine, cystatin C, and SAPS-3 for predicting AKI, RRT, and 30-day mortality are presented in Table 3.

**Table 3.** Performance of endostatin and other variables upon ICU admission for predicting AKI, RRT, and 30-day mortality in the general ICU study population and the sepsis subgroup. *AUC* area under curve, *CI* confidence interval, *ICU* intensive care unit, *AKI* acute kidney injury, *RRT* renal replacement therapy. *SAPS-3* Simplified Acute Physiology Score 3.

| Outcome & Predictor(s) | AUC (95% CI) |  | Brier Score |  |
| --- | --- | --- | --- | --- |
|  | General ICU | Sepsis | General ICU | Sepsis |
| **Future AKI** |  |  |  |  |
| Endostatin | 0.52 (0.50-0.54) | 0.56 (0.52-0.59) | 0.19 | 0.17 |
| Creatinine | 0.67 (0.65-0.69) | 0.72 (0.69-0.75) | 0.18 | 0.16 |
| Cystatin C | 0.63 (0.61-0.65) | 0.68 (0.65-0.71) | 0.19 | 0.16 |
| Endostatin, creatinine | 0.69 (0.67-0.71) | 0.73 (0.70-0.76) | 0.18 | 0.16 |
| Endostatin, creatinine, cystatin C | 0.69 (0.68-0.71) | 0.73 (0.70-0.76) | 0.18 | 0.16 |
| **Future AKI stage 3** |  |  |  |  |
| Endostatin | 0.66 (0.64-0.68) | 0.65 (0.62-0.68) | 0.16 | 0.18 |
| Creatinine | 0.66 (0.64-0.68) | 0.63 (0.60-0.66) | 0.17 | 0.18 |
| Cystatin C | 0.67 (0.65-0.69) | 0.64 (0.61-0.67) | 0.17 | 0.18 |
| Endostatin, creatinine | 0.65 (0.63-0.68) | 0.65 (0.61-0.68) | 0.16 | 0.17 |
| Endostatin, creatinine, cystatin C | 0.68 (0.63-0.70) | 0.67 (0.64-0.70) | 0.16 | 0.17 |
| **RRT** |  |  |  |  |
| Endostatin | 0.76 (0.74-0.78) | 0.75 (0.72-0.77) | 0.11 | 0.13 |
| Creatinine | 0.84 (0.82-0.86) | 0.83 (0.80-0.85) | 0.096 | 0.13 |
| Cystatin C | 0.82 (0.81-0.84) | 0.80 (0.78-0.83) | 0.097 | 0.13 |
| Endostatin, creatinine | 0.84 (0.82-0.85) | 0.82 (0.80-0.85) | 0.096 | 0.12 |
| Endostatin, creatinine, cystatin C | 0.84 (0.83-0.86) | 0.82 (0.80-0.85) | 0.094 | 0.12 |
| **30-day mortality** |  |  |  |  |
| Endostatin | 0.60 (0.58-0.62) | 0.56 (0.54-0.59) | 0.19 | 0.20 |
| SAPS-3 | 0.78 (0.77-0.80) | 0.73 (0.71-0.75) | 0.16 | 0.18 |
| Endostatin, SAPS-3 | 0.78 (0.77-0.80) | 0.73 (0.71-0.75) | 0.16 | 0.18 |
| Endostatin, age | 0.65 (0.63-0.66) | 0.62 (0.59-0.65) | 0.18 | 0.19 |

#### Association with AKI

Adjusted ORs were 1.6 (95% CI 1.4-1.7) for endostatin, 3.2 (95% CI 2.7-3.8) for creatinine, and 1.5 (95% CI 1.3-1.7) for cystatin C.

#### Prediction of future AKI

Adjusted ORs were 1.5 (95% CI 1.4-1.7) for endostatin, 0.46 (95% CI 0.40-0.52) for creatinine, and 0.85 (95% CI 0.74-0.98) for cystatin C. Endostatin had inferior discrimination compared to creatinine (AUC 0.52 vs. 0.67, p<0.001) and cystatin C (AUC 0.63, p<0.001). Adding endostatin to creatinine improved AUC compared to creatinine alone (0.69 vs. 0.67, p=0.0016).

In the sepsis subgroup, adjusted ORs were 1.5 (95% CI 1.3-1.8) for endostatin, 0.43 (95% 0.34-0.53) for creatinine, and 0.76 (95% CI 0.61-0.96) for cystatin C. Endostatin was inferior to creatinine (AUC 0.56 vs. 0.72, p<0.001) and cystatin C (AUC 0.56 vs. 0.68, p<0.001). Adding endostatin to creatinine did not improve AUC compared to creatinine alone (0.73 vs. 0.72, p=0.079).

In admissions with a creatinine <100 μmol/L on ICU admission, endostatin (AUC 0.62, 95% CI 0.59-0.65) outperformed creatinine (AUC 0.51, 95% CI 0.49-0.54) and cystatin C (AUC 0.53, 95% CI 0.50-0.56) (both p<0.001).

#### Prediction of future AKI stage 3

Adjusted ORs were 1.4 (95% 1.2-1.6) for endostatin, 0.78 (95% CI 0.67-0.88) for creatinine, and 1.7 (95% CI 1.4-2.0) for cystatin C. Endostatin had similar discrimination to creatinine (AUC 0.66 vs. 0.66, p=0.50) and cystatin C (AUC 0.66 vs. 0.67, p=0.20). Adding endostatin to creatinine did not improve AUC (0.66 vs. 0.65, p=0.63) compared to creatinine alone.

In the sepsis subgroup, adjusted ORs were 1.4 (95% CI 1.2-1.6) for endostatin, 0.83 (95% CI 0.68-1.0) for creatinine, and 1.5 (95% CI 1.2-1.9) for cystatin C. Endostatin had equal discrimination to creatinine (AUC 0.65 vs. 0.63, p=0.21) and cystatin C (AUC 0.65 vs. 0.64, p=0.27). Adding endostatin to creatinine did not improve AUC (0.65 vs. 0.63, p=0.27).

In admissions with a creatinine <100 μmol/L on ICU admission, endostatin (AUC 0.63, 95% CI 0.59-0.68) exhibited higher AUC than creatinine (0.57, 95% CI 0.52-0.62) (p=0.031), but not cystatin C (0.61, 95% CI 0.56-0.66) (p=0.20).

#### Prediction of RRT

Adjusted ORs were 1.2 (95% CI 1.1-1.4) for endostatin, 2.2 (95% CI 1.8-2.5) for creatinine, and 1.6 (95% CI 1.4-2.0) for cystatin C. Endostatin’s discrimination was inferior to creatinine (AUC 0.76 vs. 0.84, p<0.001) and cystatin C (AUC 0.76 vs. 0.82, p<0.001). Adding endostatin to creatinine did not enhance prediction (AUC 0.84 vs. 0.84, p=0.47).

In the sepsis subgroup, adjusted ORs were 1.2 (95% CI 1.05-1.5) for endostatin, 2.3 (95% CI 1.9-2.9) for creatinine, and 1.5 (95% CI 1.2-2.0) for cystatin C. Endostatin was inferior to creatinine (AUC 0.75 vs. 0.83, p<0.001) and cystatin C (AUC 0.75 vs. 0.80, p<0.001). Adding endostatin to creatinine did not improve prediction compared to creatinine alone (AUC 0.83 vs. 0.82, p=0.66).

In admissions with a creatinine <100 μmol/L on ICU admission, endostatin (AUC 0.68, 95% CI 0.62-0.75) had better discrimination than creatinine (AUC 0.58, 95% CI 0.50-0.65) (p=0.023), but not cystatin C (AUC 0.64, 95% CI 0.56-0.71) (p=0.15).

#### Prediction of 30-day mortality

Adjusted OR was 1.0 (95% CI 0.96-1.1) for endostatin. Endostatin was inferior to SAPS-3 (AUC 0.60 vs. 0.78, p<0.001). Adding endostatin to SAPS-3 did not improve AUC (0.78 vs. 0.78, p=0.28). Combining endostatin with age was also inferior to SAPS-3 (AUC 0.65 vs. 0.78, p<0.001).

In the sepsis subgroup, the adjusted OR was 0.99 (95% CI 0.88-1.1) for endostatin. Endostatin was inferior to SAPS-3 (AUC 0.56 vs. 0.73, p<0.001). Adding endostatin to SAPS-3 did not improve AUC (0.73 vs. 0.73, p=0.29). Combining endostatin with age was also inferior to SAPS-3 (AUC 0.62 vs. 0.73, p<0.001).

#### Prediction of time to AKI

HRs were 1.2 (95% 1.1-1.2) for endostatin, 0.96 (95% 0.91-1.0) for creatinine, and 1.4 (95% 1.3-1.5) for cystatin C.

## Discussion

In this large ICU cohort, endostatin upon ICU admission was associated with AKI, future AKI, future AKI stage 3, and RRT independently of creatinine and cystatin C. In sepsis, endostatin was associated with future AKI, future AKI stage 3, and RRT independently of creatinine and cystatin C. Adding endostatin to creatinine improved the prediction of future AKI in a general ICU population but not in sepsis. In admissions with low to mildly elevated creatinine, endostatin was superior to creatinine and cystatin C in predicting future AKI and superior to creatinine in predicting RRT.

Endostatin was equal to creatinine and cystatin C in predicting future AKI stage 3 in both a general ICU population and sepsis. In addition, endostatin was associated with time to AKI independently of creatinine and cystatin C. These results indicate that endostatin is an early marker for AKI and RRT needs in a general ICU population and sepsis.

Creatinine did not predict future AKI in admissions with low to mildly elevated admission creatinine. This limits the value of creatinine as a biomarker for AKI in patients with low body weight and sarcopenia. Endostatin, alone or combined with other novel biomarkers, could play an important role in identifying AKI in these patients. Even though the absolute AUC for endostatin in predicting AKI in these patients was relatively moderate, it outperformed both creatinine and cystatin C. This is an important finding since this group had an AKI rate of almost 50%.

Our results also show that endostatin, both alone and combined with age, is inferior to SAPS-3 in predicting 30-day mortality, which applies both in a general ICU population and sepsis. Adding endostatin to SAPS-3 did not improve discrimination. This makes endostatin unlikely to be of value as a predictor of mortality but does not exclude that endostatin could be useful for predicting mortality in combination with other variables. Higher endostatin was associated with sepsis, comorbidity, organ dysfunction, ICU length of stay, and 30-day mortality.

Endostatin has previously been shown to predict AKI in intensive care in significantly smaller studies. A study from 2016 found that adding endostatin to a predictive model significantly improved AKI prediction ^21^. A more recent study with 298 patients concluded that admission endostatin levels, age, and creatinine predicted AKI development and the need for RRT in a general ICU population ^12^. Furthermore, they found that the combination of creatinine and endostatin was closely associated with AKI. These results are in line with this study.

However, a prospective multicentre study with 1112 patients from 2017 concluded that endostatin has limited value as a predictive marker for AKI and RRT, despite increasing endostatin with KDIGO stages ^22^. They found considerably lower AUC for RRT. These contrasting results compared to our study could have several explanations. Our study had a larger number of patients, a higher rate of AKI, a higher RRT rate, and higher median endostatin values. They had a 90-day mortality of 19.8%, compared to 26% 30-day mortality in this study. These differences in patient populations likely explain some of the discrepancy in results between the studies. Additionally, they did not perform subgroup analysis on patients based on admission creatinine or adjust for creatinine and cystatin C.

Despite the mentioned differences in patient populations, the AUC for 90-day mortality was comparable to our AUC for 30-day mortality ^22^. This strengthens the conclusion that endostatin is unlikely to be of value in mortality prediction despite some conflicting results in relatively smaller studies ^12^.

Endostatin elevation in AKI may be due to accelerated turnover of collagen XVIII, which is an integral part of the basement membranes in the kidney ^3^. In animal AKI models, upregulated renal endostatin expression has been shown to precede deteriorating kidney function by several hours ^35,36^. However, since collagen XVIII is part of the basement membranes in many body structures, endostatin is unlikely to be specific to kidney injury ^37^. It is also possible that endostatin reflects GFR, as its size (20kDa) should allow it to be relatively freely eliminated by glomerular filtration ^1,38,39^. Collagen metabolism is elevated in sepsis, which might explain why sepsis was associated with higher endostatin in our and similar studies ^22,40^. However, this could also be related to a higher AKI rate of sepsis in our population.

Cystatin C has been well-studied, but its role in AKI prediction is still debated. Cystatin C performed well in predicting AKI and RRT in this study, which is mainly in line with a meta-analysis of prospective cohort trials from 2017 ^41^. The addition of cystatin C to creatinine and endostatin improved some AKI prediction models in this study, but cystatin C did not predict future AKI in patients with low to mildly elevated creatinine.

Fair comparisons between endostatin and creatinine are difficult since most AKI classifications rely heavily on creatinine, including the KDIGO criteria used in this study ^29,42^. This applies not only to the outcome of future AKI, but also RRT, creating a biased comparison. Even though elevated or rising creatinine is not a dialysis indication, it will likely influence the decision to initiate RRT^43^. This should be considered when assessing the predictive performance of creatinine in this study. Using estimated GFR from both creatinine and cystatin C in combination with a GFR-based AKI classification could be a better option when assessing new biomarkers in the prediction of AKI ^39,44,45^. However, baseline cystatin C is seldom available in retrospective studies.

Creatinine is associated with delay in identifying renal function loss ^17,18^. Incorporating biomarkers such as endostatin into clinical practice could enhance early risk assessment and facilitate prompt identification of AKI, which is associated with substantial mortality and morbidity ^13,15^. Timely recognition of AKI is crucial for mitigating factors such as hypoperfusion, nephrotoxic medications, and other associated risk factors, thereby potentially improving patient outcomes and reducing the burden of AKI-related complications in the ICU.

This is the most extensive study on endostatin in an ICU setting. It was performed in multiple centres, including university and medium-sized hospitals. We utilised a comprehensive approach to data collection, including prospectively collected blood samples, electronic medical records, and registry data. This allowed for various clinical parameters in addition to SAPS-3 and SOFA. Endostatin was compared to creatinine and cystatin C, with very few missing values. Additionally, the size of the study facilitated subgroup analyses, enabling endostatin to be assessed in clinically relevant subgroups.

Prospective validation of endostatin in predicting AKI and RRT, especially in patients with low to mildly elevated initial creatinine, is warranted. Future studies exploring the integration of endostatin with other emerging AKI biomarkers would be of interest.

### Limitations

While we utilised robust methodologies to mitigate biases, the retrospective design of this study was still subject to limitations such as incomplete data. Comorbidities not included in SAPS-3 and some biochemistry (CRP, lactate, and procalcitonin) had high rates of missing data. Body weight was missing in many patients and needed to be imputed for KDIGO classification. Missing baseline creatinine could lead to an overestimation of AKI, as some patients with CKD might be misclassified as AKI.

The decision to exclusively consider patients with an ICU length of stay of 24 hours or more warrants discussion. While this criterion was set to filter out patients briefly monitored in the ICU, it also excludes patients who died shortly after arrival, consequently leaving the study population without the extremes of disease severity. Exclusion of admissions who died within 24 hours might have introduced some selection bias since this group most likely would have had a high AKI rate.

We chose to include admissions rather than individual patients. Some patients may have multiple ICU admissions, resulting in their presence as multiple cases in the database. While this could affect outcomes, it is important to acknowledge that AKI risk factors and criteria can change rapidly. Therefore, treating multiple admissions as distinct cases allows for the dynamic nature of AKI to be appropriately captured in our analyses.

Our study focused on endostatin upon ICU admission, providing a snapshot of its predictive value. However, endostatin may fluctuate in response to changes in patient status or treatment interventions ^23^. Future studies incorporating serial assessments of endostatin may provide further insights into its dynamics and prognostic utility.

## Conclusion

This large multicenter study demonstrates that endostatin is an early and potentially clinically useful biomarker for predicting AKI and RRT needs at ICU admission for critically ill patients, especially those with low to mildly elevated creatinine. Neither creatinine nor cystatin C predicts future AKI in patients with low to mildly elevated creatinine. The value of endostatin as a predictor of 30-day mortality is limited as it is inferior to and does not improve SAPS-3.

## Data Availability

The datasets generated and analysed during the current study are not publicly available due to limitations in the ethical approval of the study and data management policies of Region Skåne. However, they are available from the corresponding author on request.

## Declarations

### Ethics approval and consent to participate

This study was approved by the regional ethical review board in Lund, Sweden (DNR 2015/267 and 2017/802). Patients and their next-of-kin were given the option to opt out.

### Consent for Publication

Not applicable.

### Availability of data and materials

The datasets generated and analysed during the current study are not publicly available due to limitations in the ethical approval of the study and data management policies of Region Skåne. However, they are available from the corresponding author on request.

### Authors’ contributions

AF and HF designed and funded the study. HK performed all the calculations and produced all the figures and tables. HK wrote the first draft manuscript. All authors contributed to the final manuscript.

