## supplemental for "Plasma endostatin is an early creatinine independent predictor of acute kidney injury and need for renal replacement therapy in critical care"

### Supplemental material

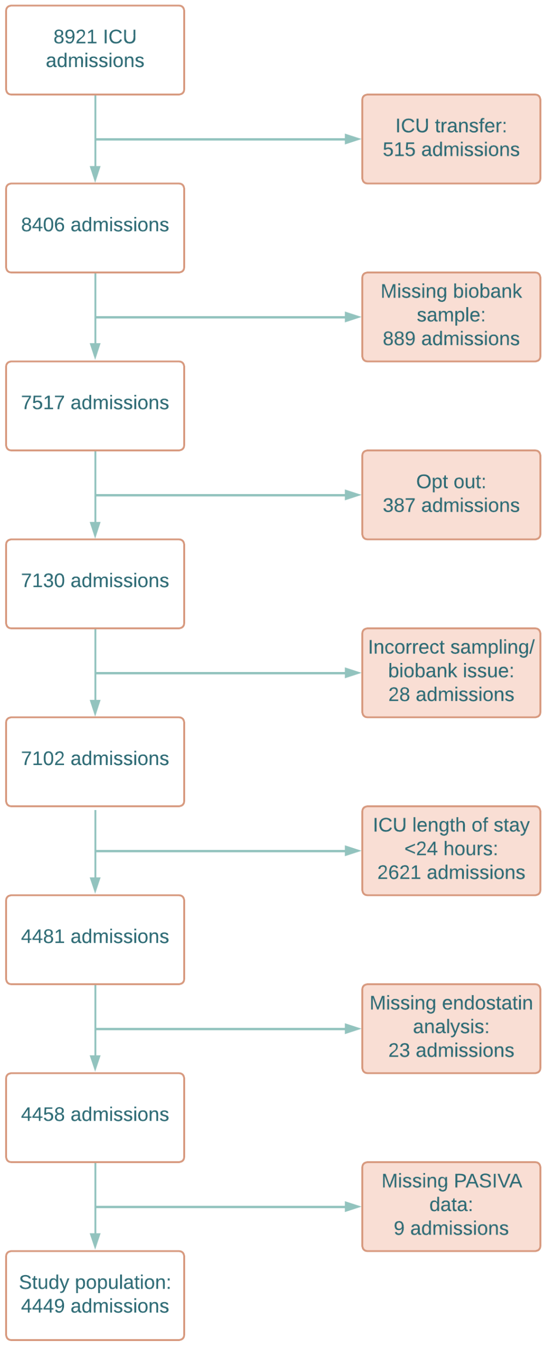


**Supplementary figure 1** Flow chart of included and excluded ICU admissions. *ICU* intensive care unit.

| | All | No AKI | AKI | p-value | Missing (%) |
|---|---|---|---|---|---|
| Number (%) | 1907 (100%) | 388 (20) | 1344 (72) | | 9.2 |
| Age (years) | 69 (59-76) | 66 (53-73) | 70 (61-77) | <0.001 | 0 |
| Male sex (%) | 61 | 57 | 62 | 0.10 | 0 |
| Body mass index (kg/m$^2$) | 27 (23-30) | 26 (22-30) | 27 (24-31) | <0.001 | 40 |
| Septic shock (%) | 53 | 45 | 58 | <0.001 | 0 |
| **Comorbidities** | | | | | |
| Immunosuppressive therapy (%) | 7.4 | 4.4 | 8.2 | 0.15 | 0 |
| Metastatic cancer (%) | 9.9 | 11 | 9.4 | 0.55 | 0 |
| Haematological cancer (%) | 4.2 | 3.6 | 4.3 | 0.64 | 0 |
| Cirrhosis (%) | 2.1 | 0.52 | 2.8 | 0.015 | 0 |
| Chronic heart failure (%) | 7.4 | 4.1 | 8.6 | 0.0051 | 0 |
| Hypertension (%) | 30 | 24 | 34 | <0.001 | 2.7 |
| Diabetes mellitus (%) | 24 | 15 | 27 | <0.001 | 4.0 |
| Ischaemic heart disease (%) | 18 | 13 | 19 | 0.0048 | 4.0 |
| Peripheral vascular disease (%) | 6.7 | 4.1 | 8.0 | 0.015 | 4.0 |
| Stroke/TIA (%) | 11 | 12 | 12 | 1.0 | 4.0 |
| Atrial fibrillation (%) | 17 | 14 | 20 | 0.018 | 4.0 |
| Asthma/COPD (%) | 17 | 20 | 16 | 0.070 | 4.0 |
| Active smoking (%) | 13 | 21 | 19 | 0.46 | 37 |
| Chronic kidney disease (%) | 11 | 2.5 | 14 | <0.001 | 4.0 |
| Chronic dialysis (%) | 2.7 | 0 | 3.9 | <0.001 | 4.0 |
| **Organ dysfunction and illness severity** | | | | | |
| SAPS-3 score | 67 (58-68) | 61 (53-70) | 70 (61-79) | <0.001 | 0 |
| SOFA score* | 8.0 (5.0-11) | 6 (4-8) | 9 (6-12) | <0.001 | 0 |
| $PaO_2/FiO_2$ (kPa) | 23 (15-37) | 23 (14-34) | 23 (15-38) | 0.35 | 9.0 |
| Cardiovascular SOFA* | 2.3 (1.7) | 1.9 (1.7) | 2.6 (1.6) | <0.001 | 0.68 |
| Mean arterial pressure (mmHg) | 65 (55-73) | 68 (60-80) | 61 (50-70) | <0.001 | 1.5 |
| Glasgow Coma Scale* | 12 (4) | 12 (4) | 12 (4) | 0.89 | 0.83 |
| Body temperature (°C) | 37 (37-38) | 38 (37-38) | 37 (36-38) | <0.001 | 0.58 |
| **Biochemistry** | | | | | |
| Plasma Endostatin (ng/mL) | 67 (50-97) | 51 (40-66) | 78 (57-110) | <0.001 | 0 |
| Baseline creatinine ($\mu$mol/L) | 80 (62-110) | 69 (54-88) | 85 (64-122) | <0.001 | 31 |
| Creatinine ($\mu$mol/L) | 115 (76-190) | 70 (55-89) | 150 (100-240) | <0.001 | 0 |
| Cystatin C (mg/L) | 1.4 (0.89-2.2) | 0.85 (0.65-1.2) | 1.8 (1.2-2.6) | <0.001 | 0 |
| White blood cell count (x$10^9$/L) | 14 (8.2-20) | 13 (8.9-18) | 14 (8.2-20) | 0.99 | 4.4 |
| Platelet count (x$10^9$/L) | 210 (140-290) | 220 (160-300) | 200 (130-290) | <0.001 | 5.6 |
| Lactate (mmol/L) | 2.2 (1.2-4.2) | 1.7 (1.0-2.7) | 2.5 (1.4-4.9) | <0.001 | 0.21 |
| C-reactive protein (mg/L) | 100 (31-220) | 100 (30-219) | 110 (37-230) | 0.11 | 0.52 |
| Procalcitonin ($\mu$g/L) | 3.2 (0.57-22) | 1.1 (0.24-5.9) | 5.3 (0.89-34) | <0.001 | 36 |
| Bilirubin ($\mu$mol/L) | 11 (7-20) | 10 (6-15) | 12 (7-22) | <0.001 | 6.0 |
| **Outcomes** | | | | | |
| Mechanical ventilation/NIV (%) | 37 | 70 | 58 | <0.001 | 4.8 |
| Vasopressor therapy (%) | 66 | 56 | 72 | <0.001 | 0 |
| CRRT (%) | 19 | 0 | 27 | <0.001 | 0 |
| ICU length of stay (days) | 3.2 (1.9-6.1) | 3.2 (2.3-5.8) | 3.7 (2.0-6.8) | 0.56 | 0 |
| 30-day mortality (%) | 28 | 23 | 31 | 0.0015 | 0 |

**Supplementary table 1** Characteristics of the sepsis subgroup. Values are medians with interquartile ranges unless otherwise specified. P-values were calculated using the Wilcoxon rank-sum test and the Pearson's chi-squared test as appropriate unless otherwise specified. *Presented as means with standard deviations, and p-value calculated using unpaired t-test. *AKI* acute kidney injury, *TIA* transient ischaemic attack, *COPD* chronic obstructive pulmonary disease, *SAPS-3* Simplified Acute Physiology Score 3, *$PaO_2$* arterial partial pressure of oxygen, *$FiO_2$* fraction of inspired oxygen (%), *SOFA* Sequential Organ Failure Assessment, *NIV* non-invasive ventilation, *RRT* renal replacement therapy, *ICU* intensive care unit.
